# Vaccine-induced antibody and T cell responses in children with acute lymphoblastic leukemia

**DOI:** 10.64898/2026.04.10.26350531

**Authors:** Janna R. Shapiro, Alina D. Dorogy, Michelle Science, Sumit Gupta, Sarah Alexander, Shelly Bolotin, Tania H. Watts

## Abstract

Children with acute lymphoblastic leukemia (ALL) are treated with multiagent chemotherapy that causes profound changes to the immune system. There are limited data on how disease and therapy impact antigen-specific immune memory, leading to inconsistent guidelines on best practices for revaccination of this population. Here, to inform vaccine guidance, we investigated whether immunity derived from routine childhood measles and varicella zoster virus (VZV) vaccines is maintained during and after therapy for childhood ALL. We report that antibodies against measles and VZV were significantly reduced in children with ALL (n=45) compared to healthy controls (n=13), particularly in older children in whom a longer time had passed since their most recent vaccine dose. However, the avidity of the measles and VZV-specific antibodies was indistinguishable between groups. Despite changes to the composition of the T cell compartment, both overall and antigen-specific T cell function were preserved in children with ALL. These data provide compelling evidence for revaccination of children following ALL treatment. Intact T cell responses suggest that post-treatment revaccination would be effective.

## Introduction

Acute lymphoblastic leukemia (ALL) is the most common childhood cancer.^1^ Pediatric ALL patients are generally treated with multiagent chemotherapy, consisting of 10-12 months of induction therapy followed by 18-30 months of maintenance therapy.^2^ Chemotherapy causes profound changes to the immune system, particularly affecting B cells and naïve proliferating cells.^3–8^ Antigen-specific humoral immunity following chemotherapy has been investigated in small cohorts with variable results, but little is known about T cell responses, which are key for reducing disease severity.^3,9–15^ This paucity of data has resulted in inconsistent guidelines and practices regarding whether children who were vaccinated prior to diagnosis should be revaccinated following treatment.^9,16,17^ North American guidelines from the Children’s Oncology Group cite the lack of evidence for their recommendation that decisions be made on an individual basis,^3^ while European guidelines recommend revaccination following treatment completion.^18–21^

The post-COVID-19 pandemic resurgence of vaccine-preventable diseases, such as measles,^22,23^ coupled with advances in the use of immunomodulatory treatment for ALL, have created a pressing need for evidence-based vaccine recommendations for children with ALL. To this end, we performed in-depth characterization of antigen-specific antibody and T cell responses in children with ALL and healthy age-matched controls to determine whether immunity derived from routine childhood measles and varicella zoster virus (VZV) vaccines is maintained during and after therapy.

## Materials and Methods

### Recruitment

Children between 2 and 18 years of age with ALL were prospectively recruited from the Leukemia and Lymphoma Clinic at the Hospital for Sick Children (SickKids), Toronto, Canada. Patients were eligible if they were either in the active maintenance phase of chemotherapy, ≤2 years after completing chemotherapy, or ≥6 months after CAR-T cell therapy. Healthy controls were recruited from the SickKids Emergency Department if they had no evidence of an active infection and were not followed by a specialist for a chronic medical condition. Written informed consent and assent were obtained from participants or their caregivers, as appropriate. This study was approved by and the Research Ethics Board of the Hospital for Sick Children (REB approval # 1000080299) and the Health Sciences Research Ethics board of the University of Toronto (REB approval # 27673).

### Data and sample collection

Data on participant demographics, medical history, and infection and vaccination history were collected via interview with participants or their caregivers. For ALL patients, we extracted additional data through chart reviews. Participants were invited to upload de-identified vaccination records, which were used to verify or supplement self-reported data. Where vaccination dates were missing, we assumed measles and VZV vaccines were given according to the recommended schedule in Ontario, Canada.^24^

Peripheral blood samples were collected at a single timepoint for healthy controls and at three timepoints for ALL patients (0, 3, and 6 months from recruitment; all data shown are from the first timepoint, in order to compare with the healthy control cohort). Samples were collected in Cell Processing Tubes (BD) and plasma and peripheral blood mononuclear cells (PBMC) were isolated and cryopreserved according to the manufacturer’s instructions.

### Antibody assays

Plasma concentration and avidity of anti-measles and anti-VZV IgG were measured via enzyme-linked immunosorbent assays (ELISA). High-binding plates (STARSTEDT) were coated overnight with measles viral lysate (Microbix, 1 µg/mL) or recombinant VZV glycoprotein E (Abcam, 2 µg/mL) in carbonate-bicarbonate buffer. Plates were blocked, and heat-inactivated plasma was diluted to 1:100 or 1:50 for measles and VZV, respectively, and run in triplicate. Standard curves were generated by serially diluting the Measles WHO Third International Standard (NIBSC 97/648) or the Varicella British Working Standard (NIBSC 90/960). To measure avidity, samples were incubated in three stepwise increases of NH₄SCN. Anti-human IgG conjugated to horseradish peroxidase (ThermoFisher) was added at 1:30,000 for measles or 1:40,000 for VZV. Plates were developed with 1-Step Ultra TMB (ThermoFisher), stopped with 2M H_2_SO_4_, and optical density (OD) was measured at 450 nm (Multiskan GO, ThermoFisher). Data were background subtracted, and outliers excluded if the coefficient of variation of triplicates exceeded 20%, resulting in one triplicate being omitted from nine of 291 samples assayed via ELISA. Antibody concentrations, in milli international units per mL (mIU/mL), were interpolated using a sigmoidal four parameter logistic standard curve. For measles, antibody concentrations were divided in half based on evidence that the Third International Standard yields titers that are approximately twice as high on ELISA compared to neutralization assays.^25,26^ Antibody levels above 120 mIU/mL were considered seropositive,^27,28^ with sensitivity analyses using cut-offs of 240 and 300 mIU/mL.^29^ For VZV, 97 mIU/mL was used as a threshold for seropositivity.^30^ Calculation of the relative avidity indices (RAI), fractional RAI (FRAI), and total RAI (TRAI) are described in **Supplemental Table 1**.

### Flow cytometry

Thawed PBMC were plated at 200,000 cells per well, rested overnight, and stimulated for 24 hours with DMSO, measles or VZV viral lysates (Microbix, 10 µg/mL), with Golgi Plug (BD) added to all wells and phorbol 12-myristate 13-acetate (PMA)/ionomycin (eBioscience) added to positive control wells for the last six hours. Samples were stained for viability (eFluor 780 fixable viability dye; eBioscience) and then fixed and permeabilized (Cytofix/Cytoperm; BD). Cells were then barcoded using two amine-binding dyes in a matrix format, such that within a set of 25 samples, each had a unique fluorescent signature.^31^ Cells in a matrix were pooled together and stained (**Supplemental Table 2**). Samples were acquired on the FACS Symphony A5 SE (BD) and analyzed in FlowJo (version 10.10), and gated with the strategy shown in **Supplemental Figure 1**. Poly-functional T cells were defined as cells producing at least two cytokines. Data were background subtracted and frequencies below 0.01 were set to 0.01.

### Cytokine secretion assays

Supernatants were collected from samples stimulated for flow cytometry prior to the addition of Golgi Plug. The concentrations of IFN*γ*, IL-2, TNF, IL-10, IL-4, IL-17, granzyme A, and granzyme B were measured using the LEGENDplex Human CD8/NK kit according to the manufacturer’s instructions. Data were acquired on the BD Fortessa X-20 and analyzed in Qognit (BioLegend). Concentrations were background-subtracted, and negative values were set to half of the lower limit of detection.

### Statistical analysis

Outcomes were compared between healthy controls and each of the ALL sub-groups (maintenance, post-treatment, or post-CAR-T) using linear regression models. Models controlled for sex, with additional vaccine-specific confounders included where appropriate. Correlations between outcomes were assessed using Pearson’s correlation coefficient.

For selected outcomes, risk factor analyses were performed only among children on maintenance. Factors of interest *a priori* included age, sex, ALL type and risk categories (standard-risk B cell ALL, high- or very high-risk B cell ALL, or T cell ALL),^32^ and months on maintenance therapy. Vaccine-related factors included the number of vaccine doses received, time since vaccination, and a composite predictor of number of doses received and age (1 dose, 3-6 years of age; 2 doses, 6-9 years of age; and 2 doses, 10-17 years of age). Simple linear regressions were employed, with raw data and sub-group predicted geometric means with 95% confidence intervals shown. Multivariate adjusted models were then built, with model coefficients and 95% confidence intervals shown. All analyses were performed in Stata19.

## Data availability

Data are available upon request to the corresponding authors.

## Results

### Participant characteristics

We enrolled 58 participants, including 45 ALL patients and 13 healthy controls, with similar demographics across study groups (**Table 1**). Four patients were treated with Blinatumomab, a B cell depleting bi-specific antibody. Due to increasing use of this therapy,^33^ data from these patients are highlighted in all figures. All four CAR-T patients had received intravenous immunoglobulin (IVIG) in the six months prior to sample collection and were therefore excluded from antibody analyses, as assays would measure antibody administered in IVIG rather than immune function in these children. All participants had received at least one dose of measles- and VZV-containing vaccines, with all vaccines administered to ALL patients prior to diagnosis (**Table 2**). One ALL patient had a documented VZV infection three months prior to sample collection, and data from this patient are highlighted where appropriate.

**Table 1.**
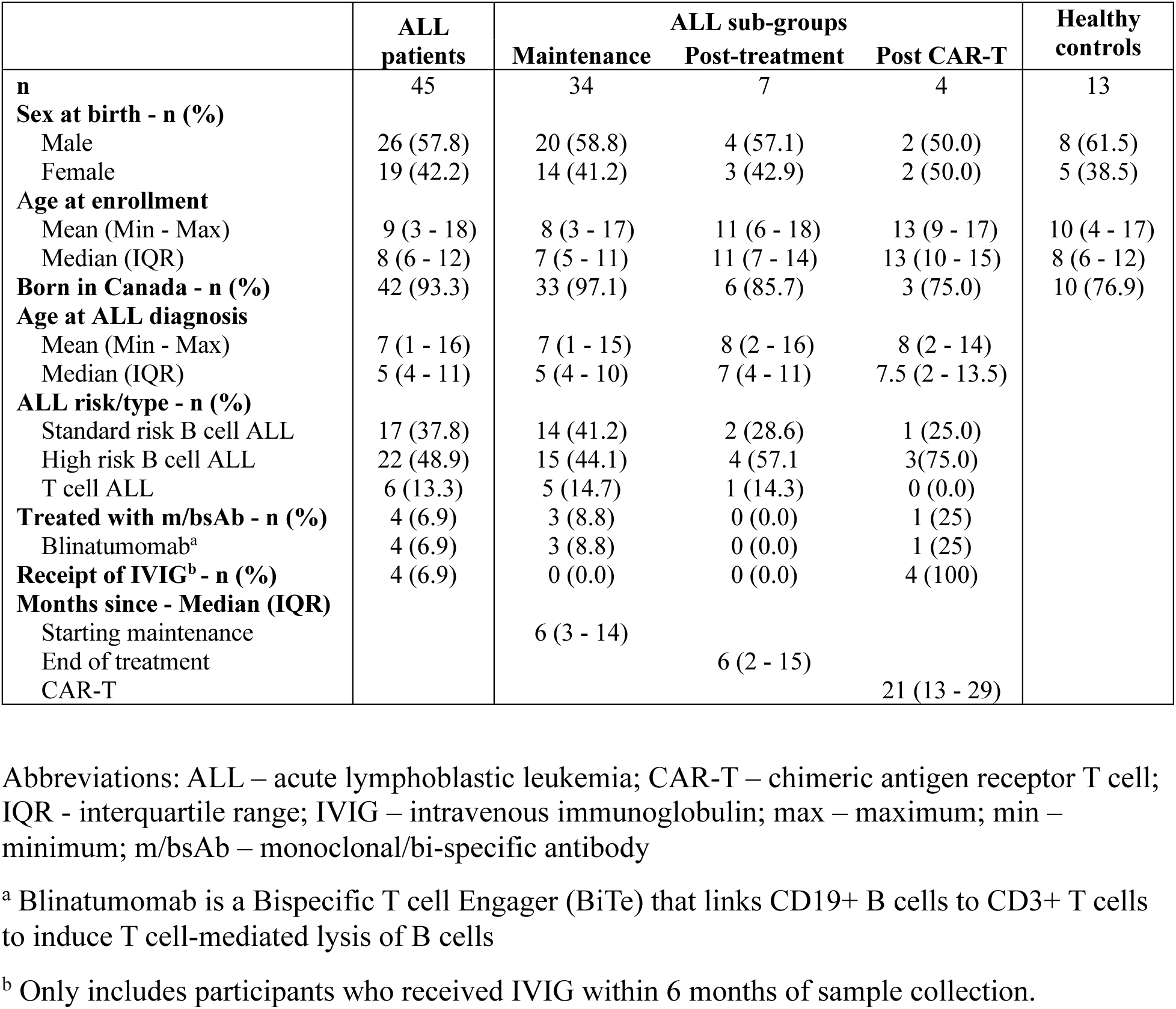
Participant characteristics.

**Table 2.**
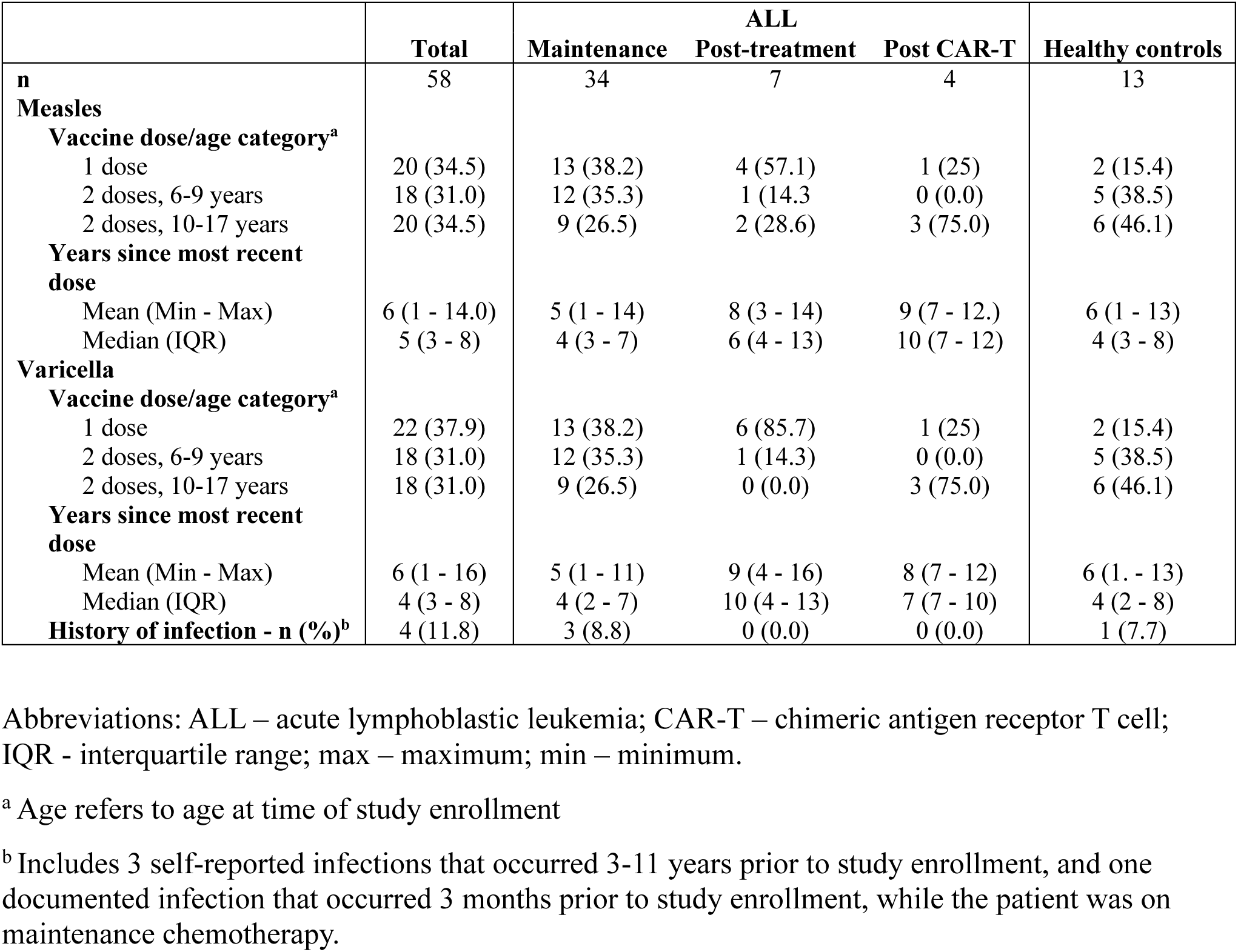
Infection and vaccination history.

Two participants were excluded from PBMC-based analyses: Insufficient blood was drawn from one healthy control, and one ALL patient on maintenance had an unusual pattern of CD3 expression, raising uncertainty as to the validity of T cell analyses (**Supplemental Figure 2**).

### Reduced antibody responses to measles vaccines administered prior to diagnosis

We first investigated the maintenance of antibodies to measles vaccines administered prior to ALL diagnosis. Children with ALL had significantly lower plasma anti-measles IgG levels relative to healthy controls when adjusting for sex and vaccine dose/age category (p<0.001 for children on maintenance, p=0.005 for children post-treatment; **Figure 1A & Supplemental Table 3**). The proportion of participants who were seronegative, as per three commonly-cited thresholds, was also higher in children with ALL than in healthy controls (**Supplemental Table 4**). The avidity of antibodies in children with ALL was similar to those observed in healthy controls and there was a weak correlation between IgG levels and TRAI (**Figure 1B-D**).

**Figure 1.**
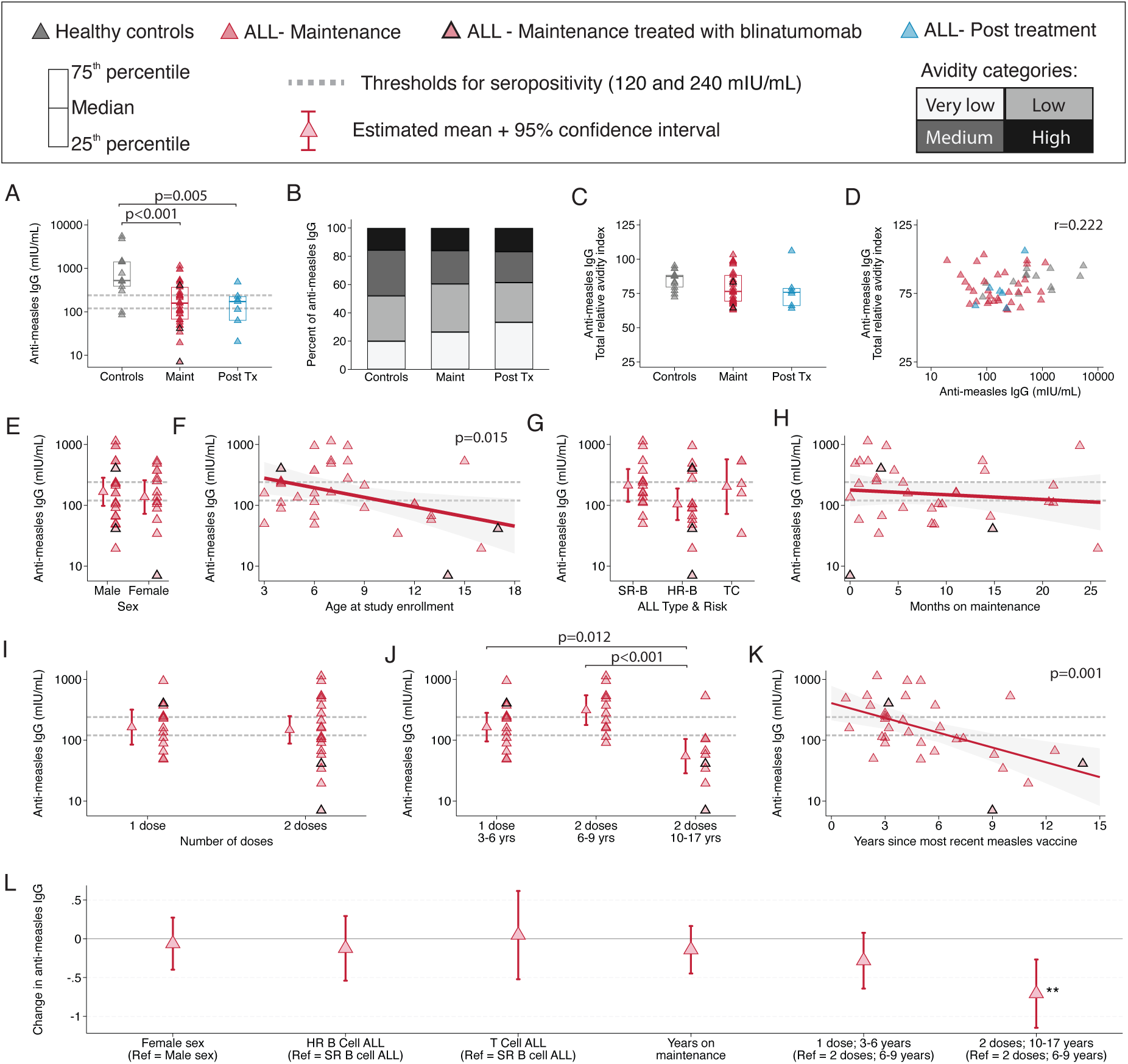
Reduced antibody responses to measles following treatment for ALL. The concentration of anti-measles plasma IgG was measured using an ELISA calibrated to the WHO Third International Standard (**A**). The ELISA was modified to assess the frequency of antibodies that were very low, low, medium, and high avidity (**B**), and avidity was summarized as the total relative avidity index (TRAI; **C**). Comparisons between groups were tested using linear regression models that controlled for sex and measles vaccine dose/age categories (**A, C**). The correlation between anti-measles IgG levels and the TRAI was assessed (**D**). Among children on maintenance chemotherapy, univariate regressions were used to assess the effects of sex (**E**), age (**F**), ALL type and risk (**G**), months on maintenance (**H**), number of measles vaccine doses (**I**), vaccine dose and age categories (**J**), and years since most recent varicella vaccine (**K**). Raw data is shown along with sub-group estimated geometric means and 95% confidence intervals (**E-K**). A multivariate regression model was then used to assess adjusted effects of the listed risk factors, with coefficients and 95% confidence intervals shown (**L**).

Focusing on children on maintenance chemotherapy, we next performed a risk factor analysis to determine whether the *a priori* factors of interest were associated with anti-measles IgG levels. While there was no significant effect of sex, ALL type and risk, or time on maintenance therapy, antibody levels decreased with age (p=0.015; **Figure 1E-H**). The number of measles vaccine doses was not significantly associated with IgG levels (**Figure 1I**). When the number of doses was incorporated with age categories, however, older two-dose vaccinated children had significantly lower antibody levels than both the one-dose (p=0.012) and younger two-dose groups (p<0.001; **Figure 1J**). Additionally, plasma anti-measles IgG levels decreased with time since receipt of a measles vaccine (p=0.001; **Figure 1K**). In the multivariate model, the only significant predictor of the concentration of anti-measles IgG was dose/age category, with the older two dose participants having predicted geometric mean levels of 119 mIU/mL (CI: 54 – 261), compared to 607 mIU/mL (CI: 336 – 1098) in the younger two dose group (p=0.003, **Figure 1L**).

### Reduced antibody responses to VZV vaccines administered prior to diagnosis

The humoral response to VZV was similar to that of measles. Children with ALL on maintenance had lower antibody levels when accounting for sex and age/vaccine dose categories (p=0.004) and higher proportions of seronegativity than healthy controls (**Figure 2A** & **Supplemental Table 4**). Antibody avidity was similar between groups, and there was no correlation between antibody levels and avidity (**Figure 2B-D**). Dynamics among children on maintenance were also similar to measles, albeit with less statistical significance (**2E-L** & **Supplemental Table 5**). Together, the measles and VZV data suggest that initial antibody responses to vaccination in children who were later diagnosed with ALL were robust, as evidenced by intact antibody avidity. Lower antibody levels and waning of responses over time, however, suggests that treatment negatively impacts antibody-producing cells.

**Figure 2.**
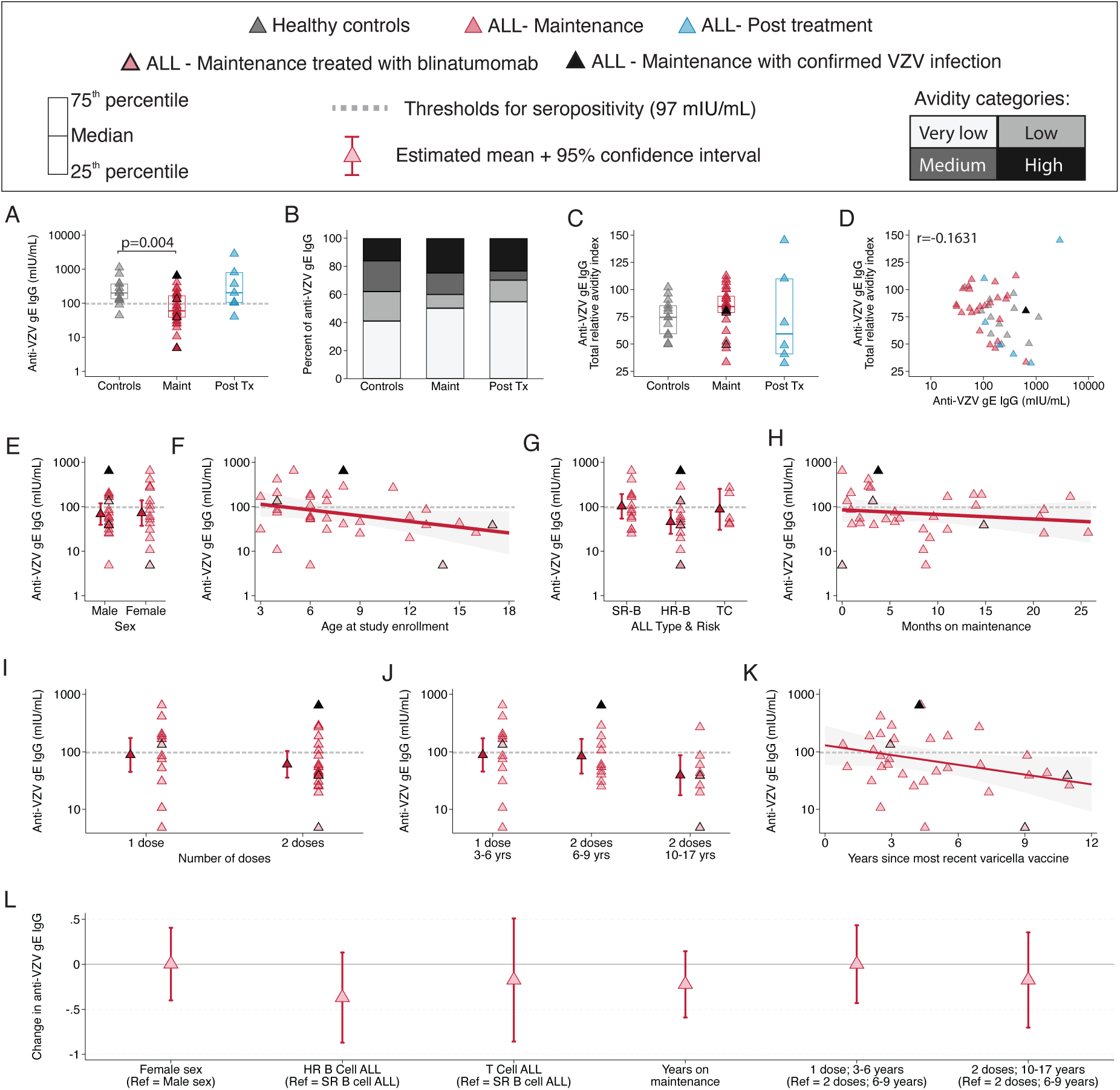
Reduced antibody responses to varicella zoster virus following treatment for ALL. Anti-varicella glycoprotein E IgG was measured using an ELISA calibrated to the Varicella British Working Standard (**A**). The ELISA was modified to assess the frequency of antibodies that were very low, low, medium, and high avidity (**B**), and avidity was summarized as the total relative avidity index (TRAI; **C**). Differences between study groups were tested using a linear regression model that controlled for sex and varicella vaccine dose/age category (**A - C**). The correlation between IgG levels and TRAI was assessed (**D**). Among children on maintenance chemotherapy, univariate regressions were used to assess the effect of sex (**E**), age (**F**), ALL type and risk (**G**), months on maintenance (**H**), number of varicella vaccine doses (**I**), vaccine dose and age categories (**J**), and years since most recent varicella vaccine (**K**). A multivariate regression model was then used to assess adjusted effects of the listed risk factors, with coefficients and 95% confidence intervals shown (**L**).

### Preservation of functional memory T cells in children treated for ALL

We next assessed the T cell compartment in children with ALL. Data abstracted from medical charts, where available, showed that children with ALL had significantly lower lymphocyte counts (**Figure 3A**). Flow cytometric analysis of unstimulated PBMC further revealed that among live lymphocytes, children on maintenance had higher frequencies of T cells relative to healthy controls (p=0.023, **Figure 3B**). Within the T cell compartment, the frequency of CD4^+^ T cells was reduced and the frequency of CD8^+^ T cells was elevated for children with ALL relative to healthy controls (**Figure 3C-E**). Few differences were observed across study groups in the frequencies of memory (CD45RA^−^CD27^+/-^ or CD45RA^+^CD27^−^) and naïve (CD45RA^+^CD27^+^) populations (**Figure 3F-I**).

**Figure 3.**
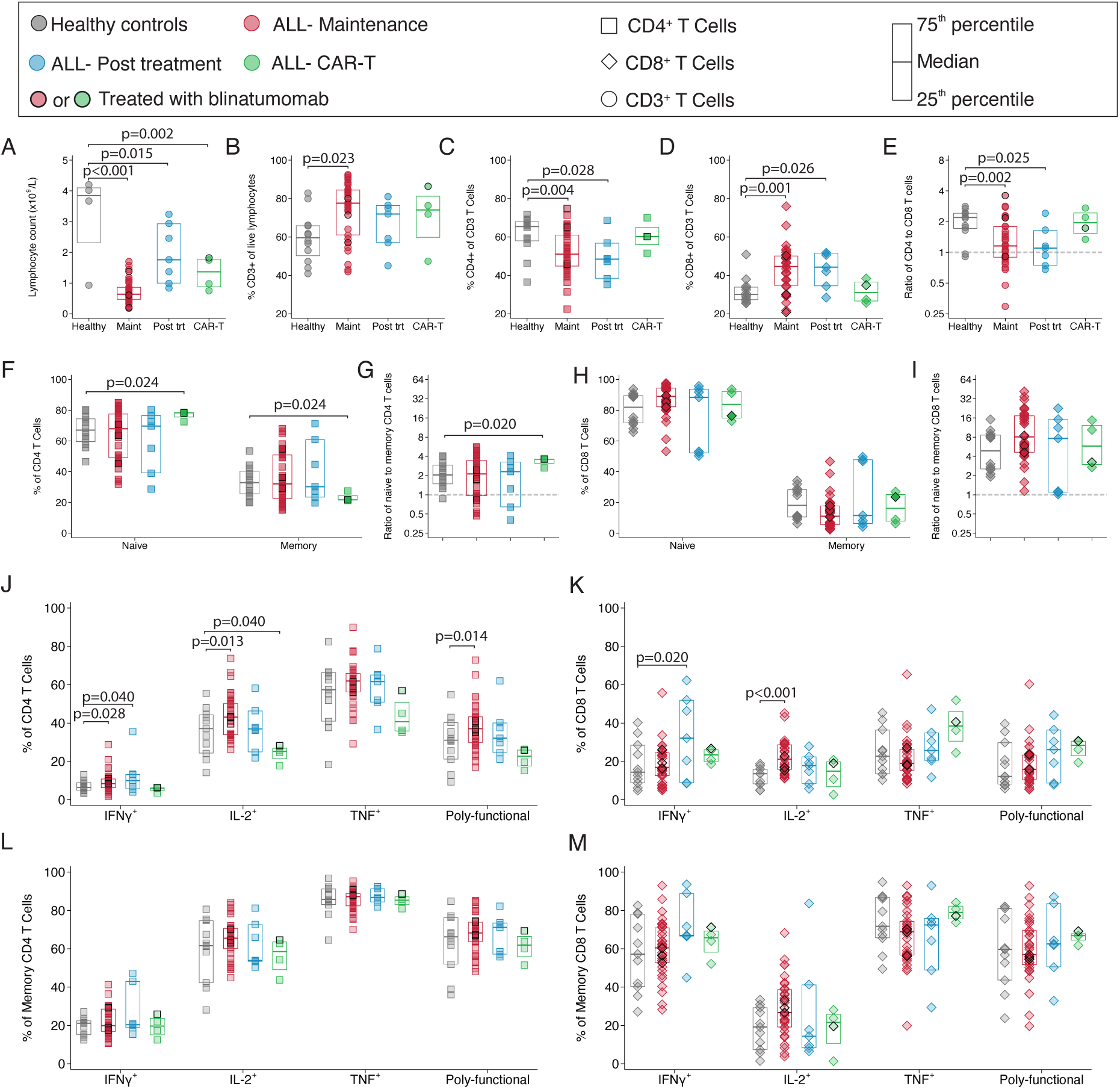
Preservation of functional memory T cells during ALL treatment. Lymphocyte counts were abstracted from the electronic medical record for the healthy controls with available data (n=4) and the ALL patients (**A**). Using flow cytometry to examine unstimulated PBMC samples, the frequencies of CD3^+^ T cells among live lymphocytes (**B**) and of CD4^+^ (**C**) and CD8^+^ (**D**) T cells among total CD3^+^ T cells, as well as the ratio of CD4:CD8 T cells (**E**), was compared between study groups. The frequencies of naïve (CD45RA^+^CD27^+^) and memory (CD45RA^−^CD27^+/-^ or CD45RA^+^CD27^−^) T cells, as well as the ratio of naïve to memory cells was assessed for CD4 (**F-G**) and CD8 (**H-I**) T cells. Using PMA/ionomycin to induce cytokine production, T cell functionality was compared between study groups by using flow cytometry to quantify the frequencies of cytokine-positive T cells among total CD4 and CD8 populations (**J-K**) and among memory CD4 and CD8 populations (**L-M**). All comparisons between study groups were assessed using linear regressions controlling for sex and age at study enrollment.

To assess overall T cell function, we stimulated PBMC with PMA/ionomycin. Among CD4^+^ and CD8^+^ T cells, there were no outcomes for which children with ALL had lower frequencies of reactive T cells than healthy controls (**Figure 3J-K**). In contrast, several populations were enriched in children on maintenance, likely explained by preferential loss of naïve lymphocytes during therapy, leading to a relatively greater frequency of memory cells.^6^ Memory T cell responses were also similar between groups (**Figure 3L-M**). Together, despite lower lymphocyte counts, functional memory T cell populations were preserved following treatment for ALL.

### Intact memory T cell responses to measles vaccines administered prior to diagnosis

To assess the maintenance of antigen-specific T cell memory to routine childhood vaccines administered prior to ALL diagnosis, we stimulated PBMC with measles viral lysate. The frequencies of measles-specific CD4^+^ and CD8^+^ T cells relative to total T cells were similar across study groups, except for higher frequencies of CD8^+^IL-2^+^ and poly-functional T cells in children on maintenance and post-treatment relative to healthy controls (**Figure 4A-B**). Alternatively, assessing the frequency of cytokine-positive T cells relative to memory T cells (**Figure 4C-D**) or total live lymphocytes (**Supplemental Figure 3A-B**) also revealed similar responses across study groups. To assess antigen-specific T cell function independent of underlying frequencies of T cell populations, we measured the concentrations of eight cytokines in culture supernatants following stimulation with measles viral lysate. Cytokine levels were comparable between study groups, with moderately elevated IFN*γ* in children on maintenance (p=0.045; **Figure 4E**). Using TNF levels as an indicator of T cell responses, both univariate and adjusted multivariate models revealed that neither demographic-, ALL treatment-, or vaccine-related factors predicted the memory T cell response to measles among children on maintenance (**Figure 4F-M**).

**Figure 4.**
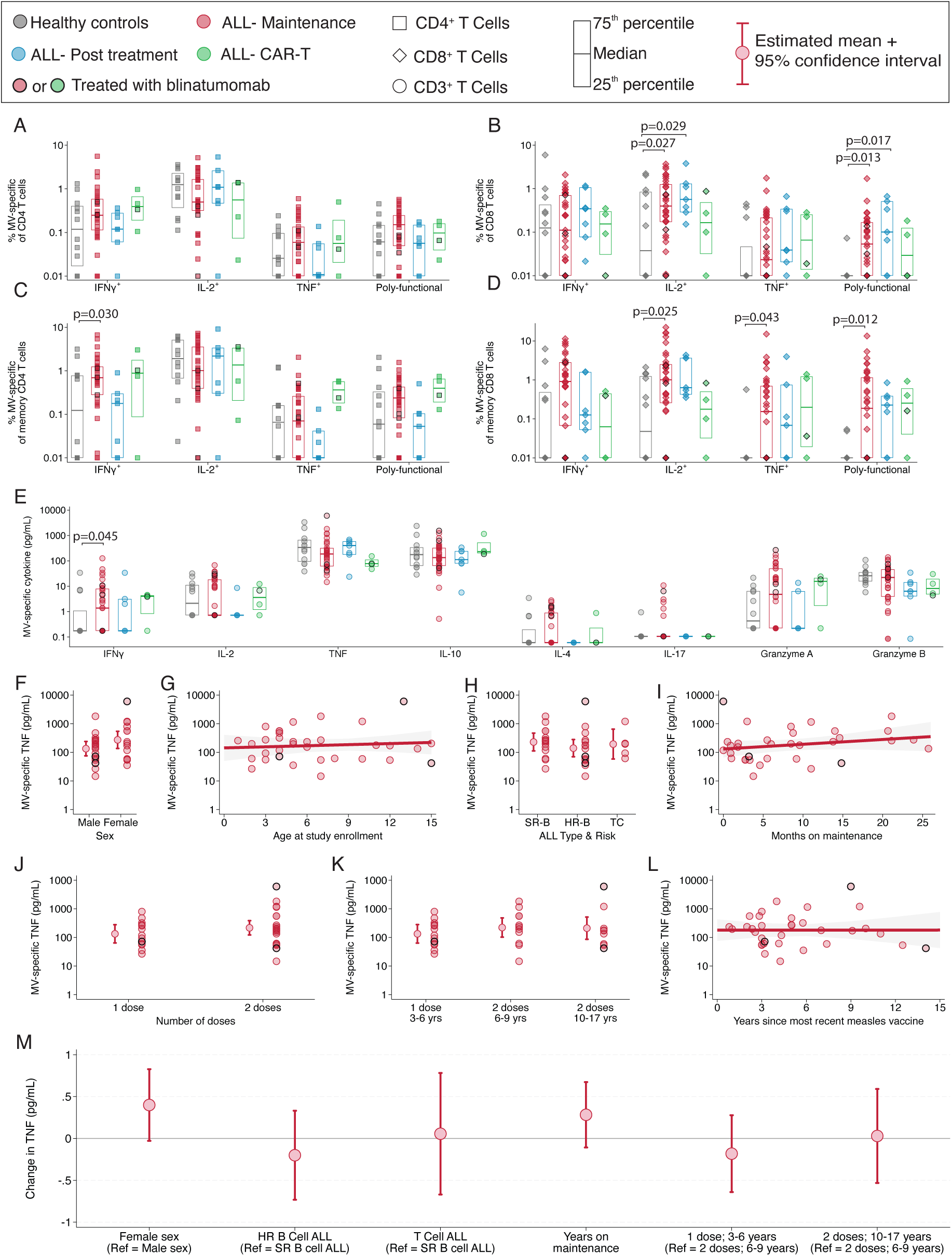
Preservation of measles specific T cell responses after ALL treatment. PBMC were stimulated with measles viral lysate and flow cytometry was used to quantify the frequency of antigen-specific CD4^+^ (**A**) and CD8^+^ (**B**) T cells among total CD4 or CD8 T cells. Responses were then re-assessed as the frequency of memory CD4 (**C**) and CD8 (**D**) T cells. The concentrations of eight secreted molecules in culture supernatants were measured using a bead-based assay and expressed as log-transformed pg/mL (**E**). Differences between groups were tested using linear regression models that controlled for sex and measles vaccine dose/age category (**A-E**). Among children on maintenance, the effects of sex (**F**), age (**G**), ALL type and risk stratification (**H**), months on maintenance (**I**), number of vaccine doses (**J**), vaccine dose and age category (**K**), and years since most recent vaccine (**L**) on the concentration of TNF were assessed using univariate regressions. A multivariate regression was then used to assess the adjusted effects of each of the listed factors on TNF concentrations among children on maintenance, with coefficients and 95% confidence intervals shown (**M**).

### Intact memory T cell responses to VZV vaccines administered prior to diagnosis

The T cell response to varicella followed a similar pattern as the response to measles, with comparable frequencies of varicella-specific cytokine-positive CD4^+^ T cells and elevated frequencies of CD8^+^ IL-2^+^ and polyfunctional T cells in children on maintenance (**Figure 5A-D & Supplemental Figure 3C-D**). Concentrations of secreted cytokines in response to antigen stimulation were also similar across study groups, except for elevated levels of granzyme A in children with ALL (**Figure 5E**). Among children on maintenance, there were no significant associations between TNF responses and any of the risk factors examined in univariate regression models (**Figure 5F-L**). In the multivariate adjusted model, however, females on maintenance had higher levels of TNF than males (p=0.035; **Figure 5M**). Taken together, we found that antigen-specific memory T cells generated by measles and VZV vaccines administered prior to ALL diagnosis were preserved during and following treatment.

**Figure 5.**
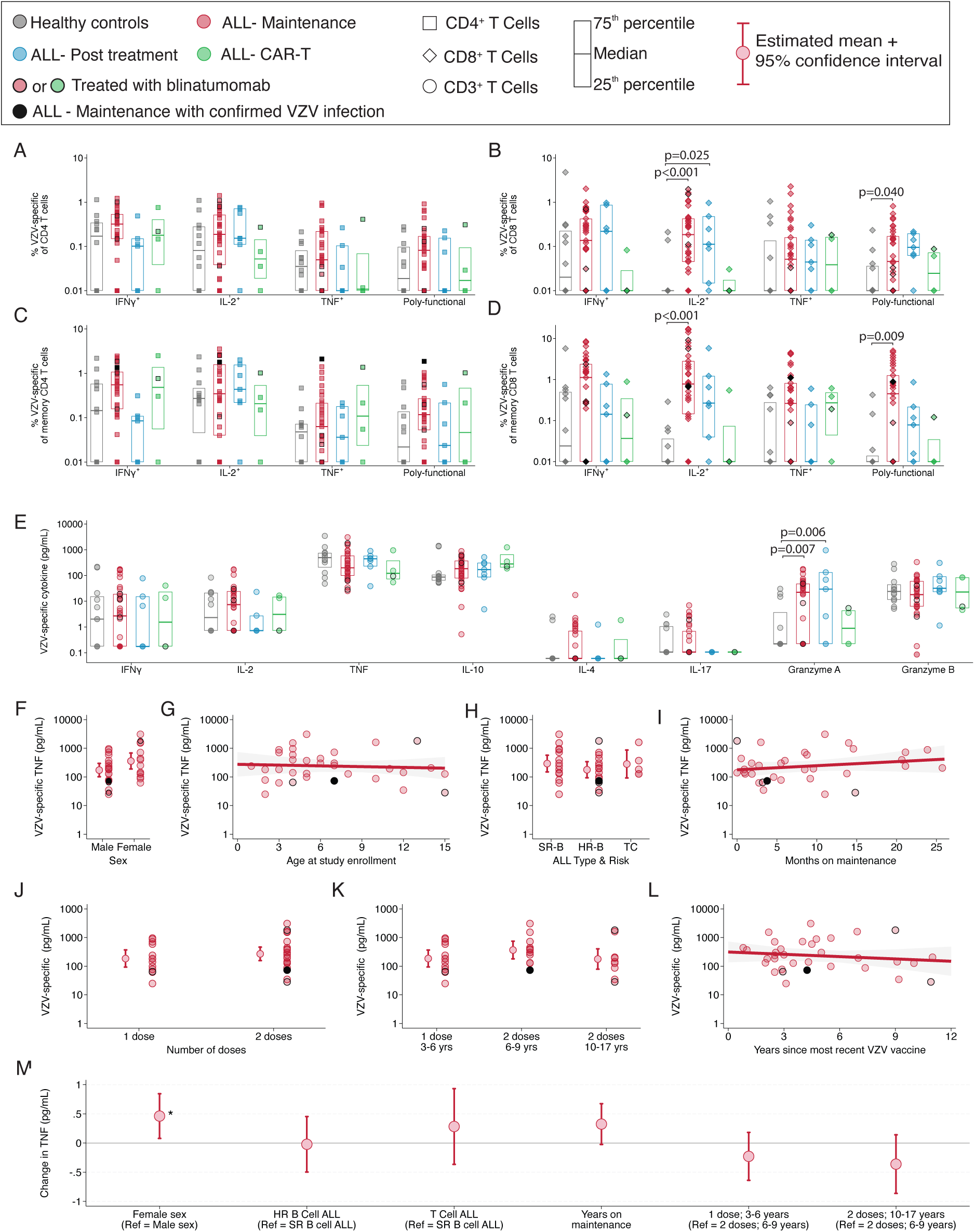
Preservation of VZV specific T cell responses after ALL treatment. PBMC were stimulated with varicella zoster viral lysate and flow cytometry was used to quantify the frequency of antigen-specific CD4 (**A**) and CD8 (**B**) T cells. Responses were then re-assessed as the frequency of memory CD4 (**C**) and CD8 (**D**) T cells. The concentrations of eight secreted molecules in culture supernatants were measured using a bead-based assay and expressed as log-transformed pg/mL (**E**). Among children on maintenance, the effects of sex (**F**), age (**G**), ALL type and risk stratification (**H**), months on maintenance (**I**), number of vaccine doses (**J**), vaccine dose and age category (**K**), and years since most recent vaccine (**L**) on the concentration of TNF were assessed using univariate regressions. A multivariate regression was then used to assess the adjusted effects of each of the listed factors on TNF concentrations among children on maintenance (**M**).

## Discussion

There is ongoing debate surrounding optimal vaccination strategies for children with ALL in the post-treatment period, resulting in confusion and discordant practices.^16,17^ As demonstrated by the COVID-19 pandemic and the post-pandemic resurgence of measles,^22,23^ there is a pressing need to generate evidence-based recommendations for revaccination in this population. This study found that children with ALL on maintenance chemotherapy or post-treatment had reduced antibody responses to vaccines administered prior to diagnosis but robust antigen-specific memory T cell responses. Together, the antibody data support the need to revaccinate children with ALL following completion of therapy, while the T cell data suggest that revaccination would be effective.

The lower vaccine-induced antibody levels in children with ALL, that decrease with both age and time since vaccination, suggest an accelerated rate of antibody waning. While some waning is expected in the years following vaccination, the rate observed in our study exceeds what has been reported in healthy children in the literature.^34^ In the years following vaccination, circulating antibody titers are maintained by long-lived plasma cells (LLPCs) that reside in the bone marrow.^35^ Evidence suggests that both the underlying leukemia and some chemotherapeutic agents can disrupt the signaling pathways that promote the survival of LLPCs,^7^ offering a potential mechanism to explain our findings. This theory aligns with the intact avidity of the remaining antibodies that we observed, as avidity is largely determined by the primary germinal center reaction that occurs when a vaccine is administered,^35^ which was several years prior to ALL diagnosis for the children in our study.

In contrast to antibodies, overall and antigen-specific T cell function were preserved in children with ALL, despite significant alterations to the composition of the T cell compartment. These findings align with previous reports on the effects of chemotherapy on various T cell subsets.^4,5,8^ Little has been published, however, on antigen-specific responses, which are key determinants of the severity of viral infections.^36–38^ The data generated here, with a several fold larger cohort than in previous studies, show that both the frequencies of memory T cells producing cytokines as well as the total amount of cytokine released in response to antigen stimulation were robust in children with ALL.

Together, our antibody and T cell data are well-supported by infectious disease epidemiology. Increased rates of vaccine-preventable infections have been observed in childhood cancer survivors,^39^ likely due to low levels of the antibodies needed to prevent infections. This underscores the pressing need to protect this population, particularly at a time when measles outbreaks are occurring globally, and endemic transmission has been re-established in the Americas two decades following measles elimination.^40^ This population, however, generally has low rates of viral infection-related mortality.^41^ The non-severe nature of viral infections are likely due to intact T cell responses, which play a critical role in resolving symptomatic disease.^36^ Our data thus support the need to revaccinate children with ALL following completion of therapy. Further, although revaccination was not directly addressed in this study, our T cell data provide a preliminary indication that if live attenuated vaccines are administered, T cell activity is likely sufficient to prevent vaccine-associated disease and support the generation of robust antibody responses.

A limitation of this study is the small sample size in the post-treatment and CAR-T groups. Therefore, more detailed data analyses were limited to the larger group of children on maintenance chemotherapy. A relatively large proportion of the children on maintenance had high-risk B cell ALL, which may not be representative of the Canadian pediatric ALL population.^42^ However, this relatively large group of high-risk children, as well as the heterogeneity of the cohort with respect to other demographic- and ALL-related factors, allowed us to thoroughly investigate how each of these factors may modify the immune outcomes of interest. In addition, in the absence of a vaccine registry in Ontario, measles and varicella vaccination dates were incomplete despite collecting vaccination records through multiple methods. To limit bias, we focused predominantly on more complete variables (i.e., number of doses and current age). Although our T cell data suggest that revaccination should be both safe and effective, further work is needed to identify optimal revaccination schedules and document vaccine efficacy and safety in this patient group. Finally, only four participants in our study were treated with Blinatumomab, which is now the standard of care for many children with B cell ALL.^43^ Of the three children on maintenance treated with Blinatumomab, one had very low antibody levels, while the other two had comparable levels to other children on maintenance. All three had similar T cell function to other children on maintenance. More work is needed to understand how Blinatumomab affects the preservation of vaccine-induced immunity.

In conclusion, our data support the need to revaccinate children with routine childhood vaccines following completion of treatment for ALL, regardless of previous vaccination history. Our T cell data explain epidemiologic findings of low levels of infection-related mortality in children with ALL. As children undergoing chemotherapy cannot receive live attenuated vaccines, our findings on the vulnerability of children with ALL to infection also reinforce the critical importance of maintaining high community vaccination coverage to protect at-risk children from vaccine-preventable diseases.

## Acknowledgements

We thank Hafsa Azher, Aarani Chandrasegaram, and Bayley Levy for their role in patient recruitment and data collection. We also thank the clinicians and staff in the Sears Cancer Clinic for their support, as well as the ALL patients and families who participated in this study.

## Author contributions

Dr. Janna Shapiro conceptualized and designed the study, performed experiments, analyzed data, and drafted the manuscript. Alina Dorogy performed experiments. Drs. Michelle Science, Sumit Gupta, and Sarah Alexander conceptualized and designed the study, coordinated and supervised participant recruitment, and critically reviewed and revised the manuscript. Dr. Shelly Bolotin and Tania Watts conceptualized and designed the study, coordinated and supervised laboratory data collection, and critically reviewed and revised the manuscript. All authors approved the final manuscript as submitted and agree to be accountable for all aspects of the work

## References

1. Ellison LF, Xie L, Sung L. Trends in paediatric cancer survival in Canada, 1992 to 2017. Health Reports 2021;32(2):3–16.

2. Inaba H, Mullighan CG. Pediatric acute lymphoblastic leukemia. Haematologica 2020;105(11):2524.

3. Guilcher GMT, Rivard L, Huang JT, Wright NAM, Anderson L, Eissa H, Pelletier W, Ramachandran S, Schechter T, Shah AJ and others. Immune function in childhood cancer survivors: a Children’s Oncology Group review. The Lancet Child & Adolescent Health 2021;5(4):284–294.

4. El-Chennawi FA, Al-Tonbary YA, Mossad YM, Ahmed MA. Immune reconstitution during maintenance therapy in children with acute lymphoblastic leukemia, relation to co-existing infection. Hematology 2008;13(4):203–209.

5. Eyrich M, Wiegering V, Lim A, Schrauder A, Winkler B, Schlegel PG. Immune function in children under chemotherapy for standard risk acute lymphoblastic leukaemia–a prospective study of 20 paediatric patients. British journal of haematology 2009;147(3):360–370.

6. Saghafian-Hedengren S, Sverremark-Ekström E, Nilsson A. T Cell Subsets During Early Life and Their Implication in the Treatment of Childhood Acute Lymphoblastic Leukemia. Frontiers in Immunology 2021;12:582539.

7. Saghafian-Hedengren S, Söderström I, Sverremark-Ekström E, Nilsson A. Insights into defective serological memory after acute lymphoblastic leukaemia treatment: The role of the plasma cell survival niche, memory B-cells and gut microbiota in vaccine responses. Blood reviews 2018;32(1):71–80.

8. Williams AP, Bate J, Brooks R, Chisholm J, Clarke SC, Dixon E, Faust SN, Galanopoulou A, Heath PT, Maishman T. Immune reconstitution in children following chemotherapy for acute leukemia. eJHaem 2020;1(1):142–151.

9. Junak SC. Lack of Consensus on Humoral Immune Status Among Survivors of Pediatric Hematological Malignancies: An Integrative Review. Journal of Pediatric Oncology Nursing 2021;38(1):51–60.

10. Patel SR, Ortín M, Cohen BJ, Borrow R, Irving D, Sheldon J, Heath PT. Revaccination of children after completion of standard chemotherapy for acute leukemia. Clinical infectious diseases 2007;44(5):635–642.

11. Top KA, Vaudry W, Morris SK, Pham-Huy A, Pernica JM, Tapiéro B, Gantt S, Price VE, Rassekh SR, Sung L. Waning vaccine immunity and vaccination responses in children treated for acute lymphoblastic leukemia: A Canadian Immunization Research Network Study. Clinical Infectious Diseases 2020;71(9):e439–e448.

12. de de la Fuente Garcia I, Coïc L, Leclerc JM, Laverdière C, Rousseau C, Ovetchkine P, Tapiéro B. Protection against vaccine preventable diseases in children treated for acute lymphoblastic leukemia. Pediatric Blood & Cancer 2017;64(2):315–320.

13. Fouda AE, Kandil SM, Boujettif F, Salama YS, Fayea NY. Humoral immune response of childhood acute lymphoblastic leukemia survivors against the measles, mumps, and rubella vaccination. Hematology 2018;23(9):590–595.

14. van Tilburg CM, Sanders EA, Rovers MM, Wolfs T, Bierings M. Loss of antibodies and response to (re-) vaccination in children after treatment for acute lymphocytic leukemia: a systematic review. Leukemia 2006;20(10):1717–1722.

15. Koochakzadeh L, Khosravi MH, Pourakbari B, Hosseinverdi S, Aghamohammadi A, Rezaei N. Assessment of Immune Response following Immunization with DTP/Td and MMR Vaccines in Children Treated for Acute Lymphoblastic Leukemia. Pediatric Hematology and Oncology 2014;31(7):656–663.

16. Top KA, Pham-Huy A, Price V, Sung L, Tran D, Vaudry W, Halperin SA, De Serres G. Immunization practices in acute lymphocytic leukemia and post-hematopoietic stem cell transplant in Canadian Pediatric Hematology/Oncology centers. Human vaccines & immunotherapeutics 2016;12(4):931–936.

17. van Thiel Berghuijs KM, Kaddas HK, Warner EL, Fair DB, Fluchel M, Knackstedt ED, Verma A, Kepka D, Green AL, Smitherman AB and others. Vaccination practices of pediatric oncologists from eight states. BMC Health Services Research 2023;23(1):1215.

18. Patel SR, Skinner R, Heath PT. Vaccinations For Paediatric Patients Treated With Standard-Dose Chemotherapy And Haemopoietic Stem Cell Transplantation (HSCT) Recipients. 2023.

19. Roussel A, Léglise C, Rialland F, Duplan M, Falaque F, Boulanger C, Cardine AM, Alimi A, Pochon C, Rabian F and others. Vaccination des enfants et adolescents traités pour une leucémie aiguë, hors allogreffés : recommandations du comité leucémies de la Société française de lutte contre les cancers et les leucémies de l’enfant et de l’adolescent (SFCE). Bulletin du Cancer 2025;112(2):208–224.

20. Cesaro S, Giacchino M, Fioredda F, Barone A, Battisti L, Bezzio S, Frenos S, Santis RD, Livadiotti S, Marinello S and others. Guidelines on Vaccinations in Paediatric Haematology and Oncology Patients. BioMed Research International 2014;2014:707691.

21. Mikulska M, Cesaro S, de Lavallade H, Di Blasi R, Einarsdottir S, Gallo G, Rieger C, Engelhard D, Lehrnbecher T, Ljungman P and others. Vaccination of patients with haematological malignancies who did not have transplantations: guidelines from the 2017 European Conference on Infections in Leukaemia (ECIL 7). The Lancet Infectious Diseases 2019;19(6):e188–e199.

22. Taylor L. Canada sees surge in measles as cases rise across North America. British Medical Journal Publishing Group; 2025.

23. Do LAH, Mulholland K. Measles 2025. N Engl J Med 2025;393(24):2447–2458.

24. Government of Ontario. 2025 17-November-2025. Ontario’s routine immunization schedule. <https://www.ontario.ca/page/ontarios-routine-immunization-schedule>. 17-November-2025.

25. Bentley M, Christian PD, Heath A. Report on collaborative study to investigate the relationship between the 1st IRP and the 2nd and 3rd International Standards for Anti-Measles Serum/Plasma, in both ELISA and PRNT. WHO Expert Committee on Biological Standardization,; 2007.

26. Bentley M, Christian PD, Heath A. Report of a Collaborative Study to Assess the Suitability of a Replacement for the 2nd International Standard for Anti-Measle Serum. 2006.

27. Bolotin S, Hughes SL, Gul N, Khan S, Rota PA, Severini A, Hahné S, Tricco A, Moss WJ, Orenstein W. What is the evidence to support a correlate of protection for measles? A systematic review. The Journal of infectious diseases 2020;221(10):1576–1583.

28. Chen RT, Markowitz LE, Albrecht P, Stewart JA, Mofenson LM, Preblud SR, Orenstein WA. Measles antibody: reevaluation of protective titers. Journal of infectious diseases 1990;162(5):1036–1042.

29. Plotkin SA, Plotkin SA. Correlates of Vaccine-Induced Immunity. Clinical Infectious Diseases 2008;47(3):401–409.

30. Cunningham AL, Heineman TC, Lal H, Godeaux O, Chlibek R, Hwang SJ, McElhaney JE, Vesikari T, Andrews C, Choi WS and others. Immune Responses to a Recombinant Glycoprotein E Herpes Zoster Vaccine in Adults Aged 50 Years or Older. J Infect Dis 2018;217(11):1750–1760.

31. Shapiro JR, Simard N, Bolotin S, Watts TH. Fluorescent Cell Barcoding of Peripheral Blood Mononuclear Cells for High-Throughput Assessment of Vaccine-Induced T Cell Responses in Low-Volume Research Samples. Cytometry Part A 2025;107(5):321–332.

32. DelRocco N, Loh M, Borowitz M, Gupta S, Rabin K, Zweidler-McKay P, Maloney K, Mattano L, Larsen E, Angiolillo A. Enhanced risk stratification for children and young adults with B-cell acute lymphoblastic leukemia: a children’s oncology group report. Leukemia 2024;38(4):720–728.

33. Gupta S, Rau RE, Kairalla JA, Rabin KR, Wang C, Angiolillo AL, Alexander S, Carroll AJ, Conway S, Gore L. Blinatumomab in standard-risk B-cell acute lymphoblastic leukemia in children. N Engl J Med 2025;392(9):875–891.

34. Bolotin S, Osman S, Hughes SL, Ariyarajah A, Tricco AC, Khan S, Li L, Johnson C, Friedman L, Gul N and others. In Elimination Settings, Measles Antibodies Wane After Vaccination but Not After Infection: A Systematic Review and Meta-Analysis The Journal of Infectious Diseases 2022;226(7):1127–1139.

35. Akkaya M, Kwak K, Pierce SK. B cell memory: building two walls of protection against pathogens. Nature Reviews Immunology 2020;20(4):229–238.

36. Shapiro JR, Corrado M, Perry J, Watts TH, Bolotin S. The contributions of T cell-mediated immunity to protection from vaccine-preventable diseases: A primer. Human Vaccines & Immunotherapeutics 2024;20(1):2395679.

37. Haining WN, Neuberg DS, Keczkemethy HL, Evans JW, Rivoli S, Gelman R, Rosenblatt HM, Shearer WT, Guenaga J, Douek DC and others. Antigen-specific T-cell memory is preserved in children treated for acute lymphoblastic leukemia. Blood 2005;106(5):1749–1754.

38. Tilburg CMv, Gent Rv, Bierings MB, Otto SA, Sanders EAM, Nibbelke EE, Gaiser JF, Janssens-Korpela PL, Wolfs TFW, Bloem AC and others. Immune reconstitution in children following chemotherapy for haematological malignancies: a long-term follow-up. Br J Haematol 2011;152(2):201–210.

39. Chehab L, Doody DR, Esbenshade AJ, Guilcher GM, Dvorak CC, Fisher BT, Mueller BA, Chow EJ, Rossoff J. A population-based study of the long-term risk of infections associated with hospitalization in childhood cancer survivors. J Clin Oncol 2023;41(2):364–372.

40. Pan American Health Organization. 2025 PAHO calls for regional action as the Americas lose measles elimination status. <https://www.paho.org/en/news/10-11-2025-paho-calls-regional-action-americas-lose-measles-elimination-status>.

41. O’Connor D, Bate J, Wade R, Clack R, Dhir S, Hough R, Vora A, Goulden N, Samarasinghe S. Infection-related mortality in children with acute lymphoblastic leukemia: an analysis of infectious deaths on UKALL2003. Blood 2014;124(7):1056–1061.

42. PDQ Pediatric Treatment Editorial Board. Childhood acute lymphoblastic leukemia treatment (PDQ®). PDQ Cancer Information Summaries [Internet]: National Cancer Institute (US); 2025.

43. Hall AG, Rau RE. Blinatumomab use in pediatric B-ALL: where are we now? Blood Advances 2025;9(15):3946–3954.

