## Supplemental material file for "Vaccine-induced antibody and T cell responses in children with acute lymphoblastic leukemia"

**Reduced vaccine-induced antibody responses and preserved T cell immunity in children with acute lymphoblastic leukemia**

**Supplemental Materials**

### Supplemental Tables

#### Supplemental Table 1. Calculation of avidity indices

| Outcome | Formulae |
|---|---|
| Relative avidity index (RAI)[a,b,c] | • $RAI_{2M} = OD_{2M}/OD_{0M}$<br>• $RAI_{1M} = OD_{1M}/OD_{0M}$<br>• $RAI_{0.5M} = OD_{0.5M}/OD_{0M}$ |
| Fractional relative avidity index (FRAI)d | • % high avidity ($FRAI_{2M}$) = $RAI_{2M}$<br>• % medium avidity ($FRAI_{1M}$) = $RAI_{1M} – RAI_{2M}$<br>• % low avidity ($FRAI_{0.5M}$) = $RAI_{0.5M} – RAI_{1M}$<br>• % very low avidity ($FRAI_{<0.5M}$) = 100 - $RAI_{0.5M}$ |
| Total relative avidity index (TRAI) | $2 * FRAI_2 + 1 * FRAI_1 + 0.5 * FRAI_{0.5} + 0.25 * FRAI_{<0.5}$ |

[a] All OD values were background subtracted. Samples with background subtracted $OD_{0M}$ <0.05 were excluded from avidity analyses (n = 2 for measles and n = 10 for VZV).
[b] $OD_{2M}$, $OD_{1M}$, and $OD_{0.5M}$ values below 0.05 were set to 0.025.
[c] RAI values above 100% were set to 100%
[d] FRAI values below 0% were set to 0%

#### Supplemental Table 2. Flow cytometry panel

| Marker | Fluorophore | Clone | Manufacturer |
|---|---|---|---|
| CD3 | Brilliant Violet 480 | UCHT1 | eBioscience |
| CD4 | Brilliant Ultraviolet 805 | RPA-T4 | eBioscience |
| CD8 | Brilliant Ultraviolet 395 | RPA-T8 | BD |
| CD14 | Real Blue 705 | M5E2 | BD |
| CD45RA | Brilliant Violet 711 | HI100 | Biolegend |
| CD27 | APC | M-T271 | Biolegend |
| TNF | Brilliant Violet 421 | Mab11 | eBioscience |
| IL-2 | Real Blue 780 | MQ1-17H12 | BD |
| IFN$\gamma$ | Real Yellow 586 | B27 | BD |

**Supplemental Table 3. Unadjusted antibody concentrations and avidity by study group**

| | Measles | Varicella |
|---|---|---|
| **IgG - Geometric mean (95% CI)** | | |
| Controls | 687.7 (320.6 - 1475.2) | 213.1 (125.7 - 361.1) |
| ALL Maintenance | 153.8 (102.8 - 230.1) | 69.9 (46.1 - 106.1) |
| ALL Post-treatment | 126.9 (49.6 - 324.6) | 258.5 (68.9 - 969.8) |
| **Avidity categories - Mean frequency (95% CI)** | | |
| High | | |
| Controls | 15.7 (13.2 - 18.2) | 16.1 (9.8 - 22.4) |
| ALL Maintenance | 16.0 (13.2 - 18.8) | 24.8 (19.8 - 29.8) |
| ALL Post-treatment | 16.8 (7.1 - 26.5) | 23.3 (-0.2 - 46.8) |
| Medium | | |
| Controls | 32.3 (25.0 - 39.6) | 21.8 (11.8 - 31.7) |
| ALL Maintenance | 23.6 (19.0 - 28.3) | 15.3 (7.4 - 23.2) |
| ALL Post-treatment | 21.8 (10.4 - 33.2) | 6.6 (-2.0 - 15.2) |
| Low | | |
| Controls | 32.0 (26.4 - 37.7) | 21.0 (14.5 - 27.6) |
| ALL Maintenance | 33.9 (29.9 - 38.0) | 9.7 (5.8 - 13.7) |
| ALL Post-treatment | 28.1 (11.0 - 45.2) | 15.3 (8.6 - 22.0) |
| Very low | | |
| Controls | 19.9 (15.0 - 24.9) | 41.0 (30.9 - 51.2) |
| ALL Maintenance | 26.4 (21.3 - 31.5) | 50.2 (40.8 - 59.7) |
| ALL Post-treatment | 33.3 (10.4 - 56.2) | 54.8 (23.1 - 86.5) |
| **Total relative avidity index – Mean (95% CI)** | | |
| Controls | 84.8 (80.3 - 89.2) | 74.8 (64.6 - 85.1) |
| ALL Maintenance | 79.2 (75.0 - 83.4) | 82.2 (73.8 - 90.6) |
| ALL Post-treatment | 77.8 (62.0 - 93.6) | 74.6 (28.0 - 121.2) |

**Supplemental Table 4. Seronegativity by study group**

| | Healthy controls | ALL Maintenance | ALL Post-treatment |
|---|---|---|---|
| **Measles** | | | |
| % below 120 mIU/mL | 15.4 | 44.1 | 42.9 |
| % below 240 mIU/mL | 15.4 | 64.7 | 85.7 |
| % below 300 mIU/mL | 15.4 | 70.6 | 85.7 |
| **Varicella** | | | |
| % below 97 mIU/mL | 15.4 | 61.8 | 14.3 |

**Supplemental Table 5. Seronegativity risk factor analysis among children with ALL on maintenance chemotherapy**

| | Measles | | | Varicella |
|---|---|---|---|---|
| | **% <120 mIU/mL** | **% <240 mIU/mL** | **% <300 mIU/mL** | **% <97 mIU/mL** |
| **Sex** | | | | |
| Males | 45.0 | 70.0 | 70.0 | 65.0 |
| Females | 42.9 | 57.1 | 71.4 | 57.1 |
| **ALL Type & Risk** | | | | |
| Standard risk B cell | 28.6 | 64.3 | 71.4 | 50 |
| High risk B cell | 66.7 | 66.7 | 73.3 | 73.3 |
| T cell | 20.0 | 60.0 | 60.0 | 60 |
| **Vaccine doses** | | | | |
| 1 dose | 38.5 | 69.2 | 76.9 | 46.2 |
| 2 doses | 47.6 | 61.9 | 66.7 | 71.4 |
| **Age/vaccine dose** | | | | |
| 1 dose; 3-6 years | 38.5 | 69.2 | 76.9 | 46.2 |
| 2 doses; 6-9 years | 16.7 | 41.7 | 50.0 | 58.3 |
| 2 doses; 10-17 years | 88.9 | 88.9 | 88.9 | 88.9 |

### Supplemental Figures

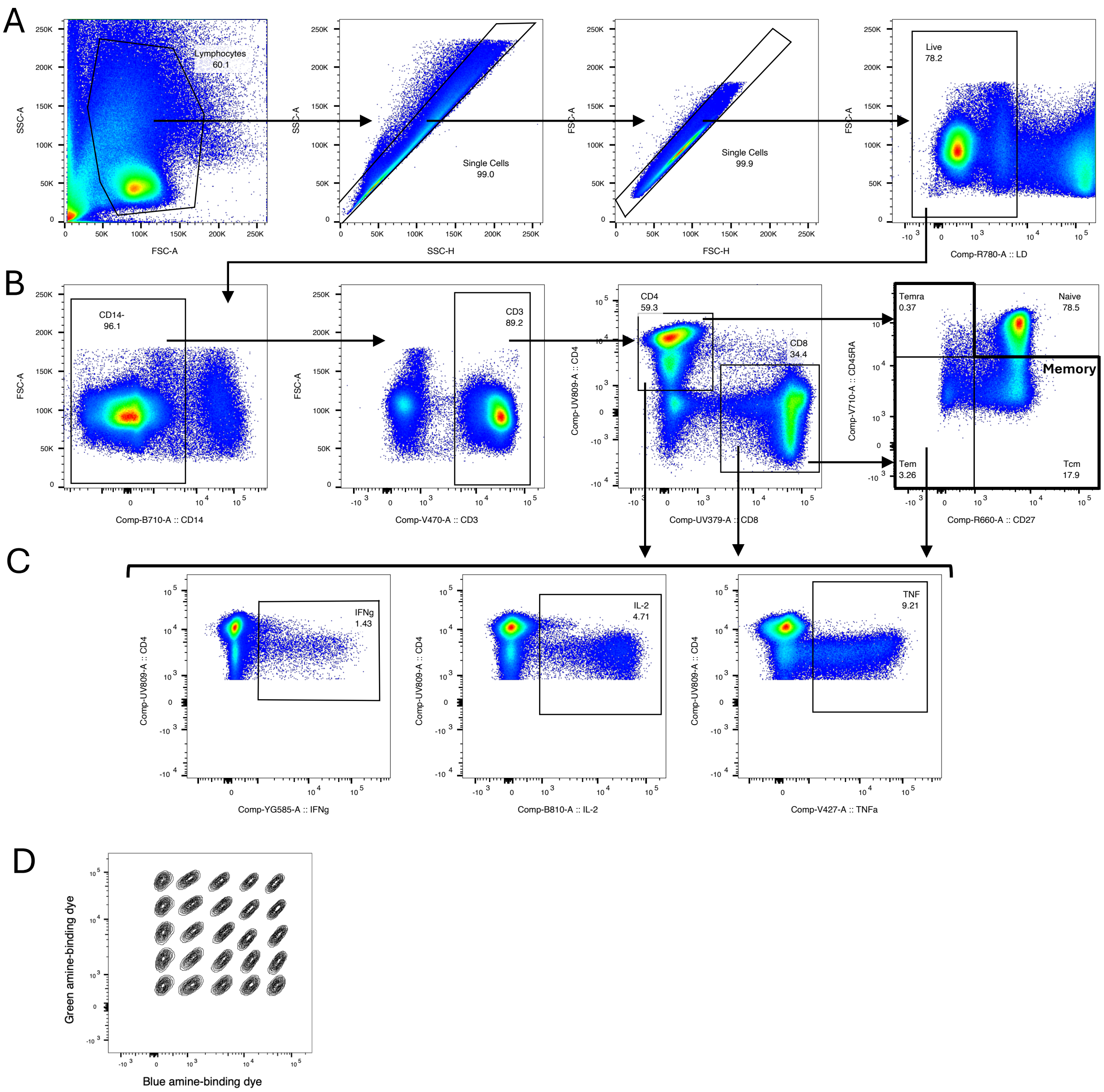

**Supplemental Figure 1. Flow cytometry gating strategy.**
Peripheral blood mononuclear cell samples were stimulated, stained for viability and then barcoded using a combination of two fixable viability dyes. Barcoded samples were pooled together and stained for extracellular phenotypic markers and intracellular cytokines. On the pooled sample, lymphocytes were gated, following by single, live cells (**A**). Among CD14$^-$ cells, CD3$^+$ T cells were gated as CD4$^+$ or CD8$^+$, and then CD45RA and CD27 were used to identify memory subsets among both CD4$^+$ and CD8$^+$ cells (**B**). Among total CD4$^+$ and CD8$^+$, as well as among memory CD4$^+$ and CD8$^+$, cytokine-positive cells were identified, and Boolean gating was used to identify poly-functional cells (i.e., cells producing multiple cytokines) (**C**). For each population of interest, the barcoding matrix was deconvoluted to calculate frequencies for each input sample (**D**).

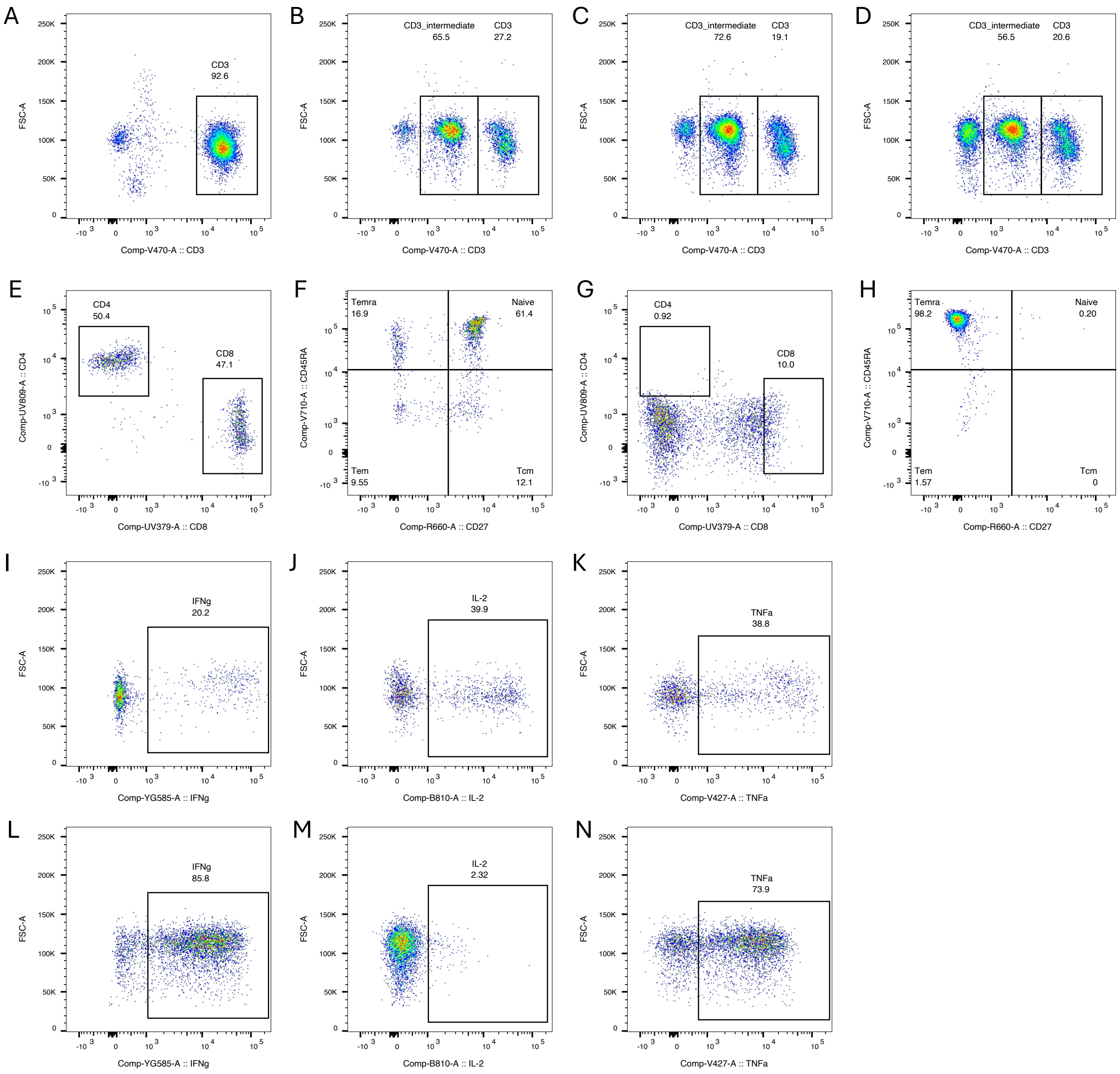

**Supplemental Figure 2. T cell phenotype from the ALL patient excluded from T cell analyses.**

In contrast to typical CD3 expression (**A**), one ALL patient had an additional T cell population with an intermediate level of CD3 expression, termed “CD3-intermediate” in samples collected a the 0- (**B**), 3- (**C**), and 6- (**C**) month timepoints. The patient was diagnosed with precursor B cell ALL at 3 years of age and was 6 years of age and on maintenance chemotherapy at the time of study enrollment. He has a diagnosis of autism spectrum disorder, but no other complications were noted in his chart. While typical CD3$^{+}$ T cells showed normal expression of CD4/CD8 (**E**) and CD45RA/CD27 (**F**), the CD3-intermediate population had no CD4 expression and down-shifted CD8 expression (**G**) and 98% were of the $T_{EMRA}$ phenotype (i.e., CD45RA$^{+}$CD27$^{+}$; **H**). Upon stimulation with PMA/ionomycin, the typical CD3$^{+}$ T cells expressed normal levels of intracellular cytokines (**I-K**), while the frequency of cytokine-positive populations were altered in in the CD3-intermediate population (**L-N**).

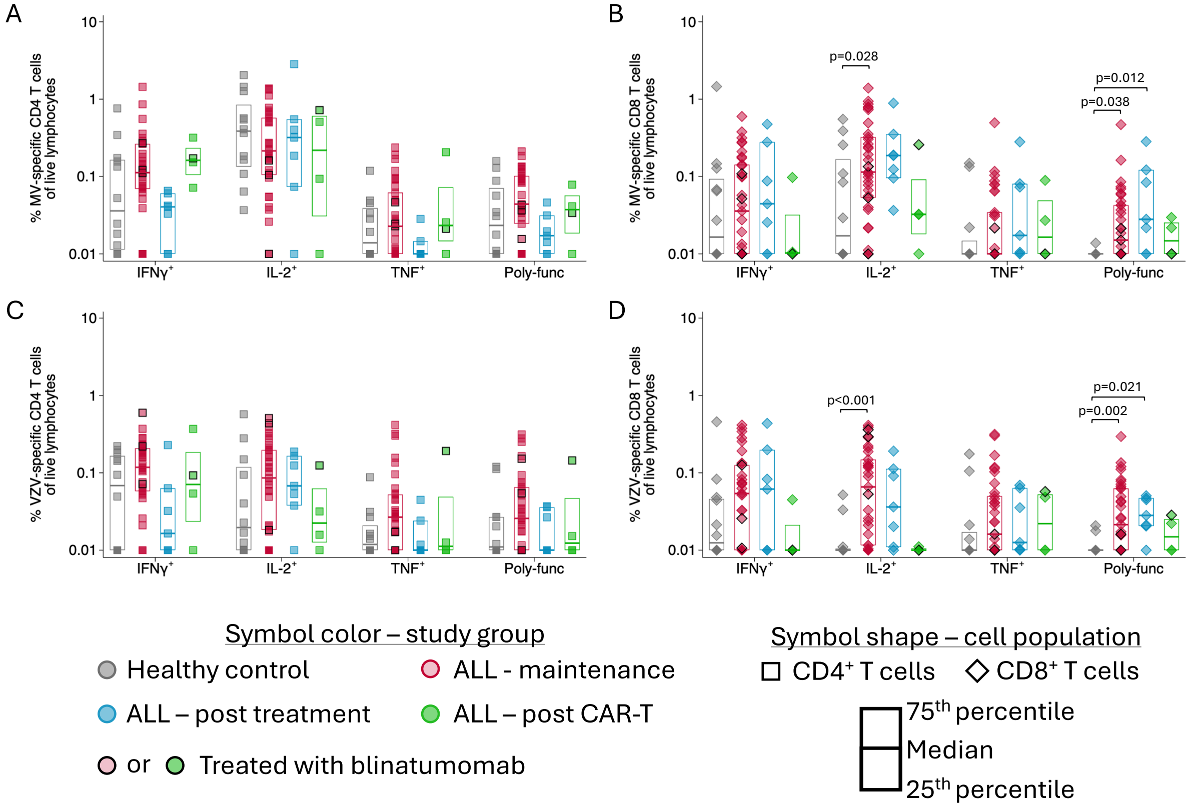

**Supplemental Figure 3. Frequency of antigen-specific T cells among live lymphocytes.** Antigen-specific T cell responses were assessed by stimulating PBMC with measles viral lysate (**A-B**), varicella zoster viral lysate (**C-D**), or SARS-CoV-2 S peptide pools (**E-F**). The frequencies of IFN$\gamma^+$, IL-2$^+$, TNF$^+$, and poly-functional CD4 and CD8 T cells were assessed as the frequency of live lymphocytes. Differences between study groups for each outcome were assessed using linear regression models that controlled for sex. For measles and varicella, models also controlled for vaccine dose/age category and years since most recent vaccine. For SARS-CoV-2, models also controlled for vaccination status and years since most recent SARS-CoV-2 exposure.
